# Estimated Veterans Health Administration costs for Alzheimer’s disease treatment with aducanumab

**DOI:** 10.1101/2021.07.24.21261063

**Authors:** Adrian D. Haimovich, Joseph L. Goulet, Cynthia A. Brandt, Asim Tarabar, Huned S Patwa, Ula Hwang

## Abstract

The approval of aducanumab for the treatment of Alzheimer’s disease has been met with significant public concern. Projections of annual cost across United States Medicare beneficiaries are in the tens of billions of dollars. Here, we performed an electronic health record review of Veterans Affairs beneficiaries, identifying 151,921 individuals (1.7%) with diagnosis codes for Alzheimer’s disease. We estimate that treatment for this cohort will cost in excess of $4 billion annually with further expenditures for associated screening, consultations, imaging, and therapeutic monitoring. The magnitude of projected expenditures necessitates early assessment of drug cost effectiveness.

## Introduction

On June 7, 2021, the United States Food and Drug Administration (FDA) approved aducanumab for the treatment of Alzheimer’s disease (AD) based on data showing reduction in amyloid plaques.^1^ This indication was later limited to mild AD. With an expected annual drug cost of $56,000, estimates of annual Medicare expenditures using census-level approximations of disease prevalence are in the tens of billions of dollars.^2^ The Veterans Health Administration (VHA) includes 1,293 health care facilities with over 9 million beneficiaries and is the largest integrated health care system in the United States. Here, we present annual cost projections for treatment with aducanumab based on a large-scale electronic health record review of Veterans Affairs (VA) patients.

## Methods

Our study included Veterans Affairs (VA) beneficiaries with at least one outpatient or inpatient visit between October 1, 2009 and September 30, 2019. VA patients with AD were identified using ICD-9 code 331.0 and ICD-10 code G30.0, 1, 8, 9. A drug price of $56,000 was used as announced by the manufacturer Biogen. Diagnostic imaging costs were determined using VA outpatient facility nationwide charges for 2020 (CPT 70551, $1,542.31). This study was approved by the VA institutional review board (1583221-6).

## Results

Of the over 9 million patient records reviewed in this study, 151,921 (1.7%) had an ICD code for AD. Prior literature suggests that approximately 50% of individuals with AD have mild disease.^3^ If half of VA patients with diagnosed AD had mild disease and were eligible for treatment with aducanumab at market cost, VA expenditures would be $4.3 billion/year, or 40% of the total VA pharmacy budget for 2021.^4^ The FDA label recommends brain magnetic resonance imaging (MRI) three times during aducanumab initiation: one within a year of starting the drug, one prior to the 7th infusion, and one prior to the 12th infusion.^5^ For this population, MRI monitoring would cost an estimated $351 million/year.

## Discussion

Patient evaluation and treatment with aducanumab poses potential significant cost for the VHA. The presented estimates are likely conservative as they do not include costs of neurologist outpatient consultation, infusions, hypersensitivity reactions, or diagnosis and management of amyloid related imaging and safety risks associated with the monoclonal antibody treatment such as microhemorrhages.^5^ Similarly, ICD codes likely underestimate disease prevalence. Population inclusion criteria may change as FDA indications are evolving.

A systematic VHA screening and treatment effort for Alzheimer’s disease could mirror the VA National Viral Hepatitis Program, which, since its inception, has cured 87% of living Veterans diagnosed with chronic hepatitis C (HCV) infection.^4^ In 2014, the FDA approved multiple direct acting antivirals for the treatment of HCV, which offered higher cure rates, improved tolerability with all-oral combination regimens, and fewer adverse events as compared to older PEG-interferon treatments^6^. While early treatment rates were limited by high costs,^6^ the VA has since treated over 128,000 patients with direct acting antivirals with a greater than 80% cure rate.^4^ Moreover, as of January 2020, 75% of all Veterans between 18 and 79 years of age have been screened for HCV.^4^ In contrast to the HCV program, aducanumab is a resource-intensive intravenous therapy that may slow cognitive decline, but is non-curative. Cost-effectiveness studies like those supporting the treatment of HCV^6^ are required to contextualize the potential longitudinal impacts on health system expenditures, which will include dementia screening, diagnostic evaluation, drug costs, and safety monitoring of treatment. These in total would threaten to overwhelm VA resources.

## Data Availability

The data presented are not publicly available.

## References

1. Office of the Commissioner. FDA Grants Accelerated Approval for Alzheimer’s Drug. Published July 6, 2021. Accessed June 25, 2021. https://www.fda.gov/news-events/press-announcements/fda-grants-accelerated-approval-alzheimers-drug

2. Cubanski J, Neuman T. FDA’s Approval of Biogen’s New Alzheimer’s Drug Has Huge Cost Implications for Medicare and Beneficiaries. Published June 10, 2021. Accessed June 25, 2021. https://www.kff.org/medicare/issue-brief/fdas-approval-of-biogens-new-alzheimers-drug-has-huge-cost-implications-for-medicare-and-beneficiaries/

3. Yuan J, Maserejian N, Liu Y, et al. Severity Distribution of Alzheimer’s Disease Dementia and Mild Cognitive Impairment in the Framingham Heart Study. J Alzheimers Dis. 2021;79(2):807–817.

4. Department of Veterans Affairs. FY 2022 Budget Submission: Medical Programs and Information Technology Programs. 2022 Congressional Submission, Volume II. Accessed July 9, 2022. https://www.va.gov/budget/docs/summary/fy2022VAbudgetVolumeIImedicalProgramsAndInformationTechnology.pdf

5. Aducanumab Label. Published 6/2021. Accessed June 25, 2021. https://www.accessdata.fda.gov/drugsatfda_docs/label/2021/761178s000lbl.pdf

6. Moon AM, Green PK, Berry K, Ioannou GN. Transformation of hepatitis C antiviral treatment in a national healthcare system following the introduction of direct antiviral agents. Aliment Pharmacol Ther. 2017;45(9):1201–1212.

